# Fibrinogen is the key factor associated with tremor relieve and recurrence in the ET patients subjected with unilateral MRgFUS thalamotomy

**DOI:** 10.1101/2024.02.14.24302845

**Authors:** Jiaji Lin, Haoxuan Lu, Dekang Zhang, Xianbing Bian, Jianxing Hu, Longsheng Pan, Xin Lou

## Abstract

**Objective:** Magnetic resonance guided focused ultrasound (MRgFUS) thalamotomy has been shown to be safe and effective to treat various types of tremor diseases, but whether the clinical status of these patients was involved was still unknown.

**Methods:** We retrospectively studied the tremor symptoms and clinical variables (at hospitalization) of 59 essential tremor (ET) patients subjected with MRgFUS thalamotomy. Patients were categorized by the short-term tremor relieve and tremor recurrence within 12 months. Multivariate logistic regression was used to screen independent factors and construct the nomograms. Additional fibrinogen knock-out mice were applied to establish ET mouse model induced by harmaline and the frequency and intensity of tremor were measured and analyzed using a force-plated accelerometer.

**Results:** It is found that 39 ET patients (66.102%) with significantly better efficiency in short-term tremor relieve (control ratio > 75%) at 1-month postoperatively, while nine ET patients (15.254%) experienced significant tremor recurrence within postoperative one-year. Multivariate analysis suggested fibrinogen (OR = 0.182, 95% CI = 0.042-0.796,) and monocyte count (OR = 0, 95% CI = 0-0.001) were independently associated with better short-term tremor relieve (>75%) at 1-month postoperatively. A total of nine ET patients (15.254%) experienced significant tremor recurrence. Admission systolic blood pressure (OR = 1.013, 95% CI = 1.010-1.062), hypertension comorbidity (OR = 2.163, 95% CI = 1.412-53.565) and fibrinogen (OR = 1.620, 95% CI = 1.047-24.376) independently contributed to tremor recurrence. The nomograms were established for better tremor relieve and tremor recurrence and have excellent performance as the AUCs were 0.829 and 0.853 (bootstrap repetition = 1000). Moreover, fibrinogen knock-out promoted a great suppression on maximum tremor motion power and overall tremor activity in ET tremor model (*P* < 0.001).

**Conclusions:** The study found that baseline fibrinogen is the key factor associated with tremor relieve and recurrence for patients subjected with MRgFUS thalamotomy. Preoperative evaluation of clinical factors has important predictive significance for short- and long-term postoperative outcomes for patients treated with MRgFUS thalamotomy.

## Introduction

Magnetic resonance guided focused ultrasound (MRgFUS) thalamotomy that targets the ventral intermedius nucleus (Vim) is a promising neuromodulation therapies and that has been shown to be safe and effective to treat various types of tremor diseases^1^. Although its application has experienced quick growth in recent years for medically refractory essential tremor (ET) and tremor-dominant Parkinson disease (PD), a growing accompanying criticism is about a variety of different efficacies (tremor improve rate ranging from 43% to 83%)^2^, with efficacy loss and tremor recurrence in long-term prognosis. In clinical trial of Zaaroor et al. for 30 patients (18 with ET, 9 with PD, and 3 with ET-PD), tremor recurring was found in 6 patients within the first 6 months^3^. In another retrospective study of 12 patients with ET, tremor recurrence in other four was noted at 1-year follow-up^4^. To sum up, it is very urgent to take these outcomes as the primary ends and investigated their relevant risk factors.

Some research has tried the optimization of technical practice of ultrasound neuromodulation to overcome such variable efficacy difference and tremor relapse, such as therapeutic parameter, targeting accuracy, lesion monitoring^5,6^. For example, there is a tendency to increase the ultrasound duration and energy threshold of MRgFUS as skull density ratio (SDR) and other factors have been found led to insufficient energy deposition as well as relatively preserved tissues^7,8^. Coordinate-based targeting techniques are often challenged by functional/tractography-based targeting techniques, which can provide more individualized targets and improve the treatment benefits^9^. However, very few studies consider potential clinical risk factors in neuromodulation therapies like MRgFUS thalamotomy and analyze their value in predicting outcomes of inefficiency and recurrence. To this end, this study retrospectively collected the clinical variables of eligible patients at hospitalization for unilateral MRgFUS thalamotomy and tried to analyze their value in predicting these therapeutic outcomes.

## Methods

### Study population and clinical variables collection

This single-center retrospective study (NCT04570046 at clinicaltrials.gov) was approved by the institutional review board and the scientific advisory committee of Chinese PLA General Hospital. All the anonymous data of patients with unilateral MRgFUS thalamotomy between 2018 to 2020 were collected in Chinese PLA General Hospital and the requirement for informed consent was waived. Medically refractory ET patients within records have received unilateral MRgFUS thalamotomy in a 3T MRI suite (Discovery 750, GE Healthcare, USA) using the ExAblate Neuro focused ultrasound system (InSightec, Israel) as previous reports^10^. Tremor performance was assessed in off-medication state using Clinical Rating Scale for Tremor (CRST) ratings preoperatively and 1/3/6/12-month postoperatively, including CRST-A assesses tremor location and amplitude, CRST-B assesses tremor when performing specific motor tasks, and CRST-C assesses functional disability due to tremor^10^. Records were excluded: 1) missing tremor assessment data at 1- or 12-month postoperatively, or less than 3 tremor assessments made within 12 months of follow-up; 2) missing values in clinical baseline variables at hospitalization; and 3) coexistence of other major diseases or undergone other brain surgeries. Finally, a total of 59 records were enrolled for present study.

The clinical variables of these patients were collected at hospitalization for MRgFUS thalamotomy, including demographic characteristics (age, gender, body mass index, admission systolic blood pressure, admission diastolic blood pressure, admission pulse rate), common comorbidity (hypertension, diabetes, hyperlipemia), hematological analysis (hemoglobin, red blood cell, white blood cell, neutrophil, lymphocyte, monocyte, eosinophil, basophil, hematocrit, mean corpuscular volume, mean corpusular hemoglobin, mean corpusular hemoglobin concerntration, red blood cell volume distribution, platelet, mean platelet volume), coagulation function (thrombin time, activated partial thromboplastin time, prothrombin time, prothrombin activity, international normalized ratio, fibrinogen), lipids Profile (cholesterol, triglyceride, apolipoprotein A1, apolipoprotein B, high-density lipoprotein cholesterol, low-density lipoprotein cholesterol, serum lipoprotein a), blood biochemistry (alanine aminotransferase, aspartate aminotransferase, total protein, serum albumin, total bilirubin, direct bilirubin, total bile acids, homocysteine, alkaline phosphatase, gamma-glutamyltransferase, glucose, urea, serum creatinine, serum uric acid, lactate dehydrogenase, total iron-binding capacity, creatine kinase, creatine kinase isoenzyme, serum calcium, inorganic phosphorus, serum magnesium, serum kalium, serum sodium, serum chloride, serum carbon dioxide, serum superoxide dismutase, phospholipid, glycosylated serum protein, serum cystatin C, serum sialic acid, glycated albumin, α-L-fucosidase, glycholic acid).

Primary hand tremor score contralateral to the Vim lesion was measured using a derived subscale composed of CRST-A and CRST-B. The relative hand tremor score ratio was calculated with the formula: postoperative scores / preoperative scores × 100%. The control ratio of hand tremor score was calculated with the formula: (postoperative scores - preoperative scores) / preoperative scores × 100%. To investigate the clinical baseline variables associated with tremor relieve and recurrence of MRgFUS thalamotomy, we defined that: (1) *short-term tremor relieve*: The tremor assessment at 1-month postoperatively was used for the measurement of short-term tremor improvement. Unilateral MRgFUS thalamotomy is considered effective with a control ratio greater than 50% at 1-month postoperatively^11^. We assigned the patients with the control ratio >75% in the better tremor relieve group to study the factors affecting effectiveness. (2) *long-term tremor recurrence*: Tremor recurrence was defined as worsening of symptoms occurring after an initial good effectiveness to MRgFUS thalamotomy with the control ratio ≥ 50% of hand tremor score at 1-month postoperatively^11^, in which hand tremor score of other postoperative timepoints (3-month, 6-month, and 12-month postoperatively) increased by 8 points and 25% compared with 1-month postoperatively. We divided the patients into the with/without tremor recurrence groups based on whether they had tremor recurrency within 12-month postoperatively. Comparisons for these ET patients were summarized in Table 1.

**Table 1.**
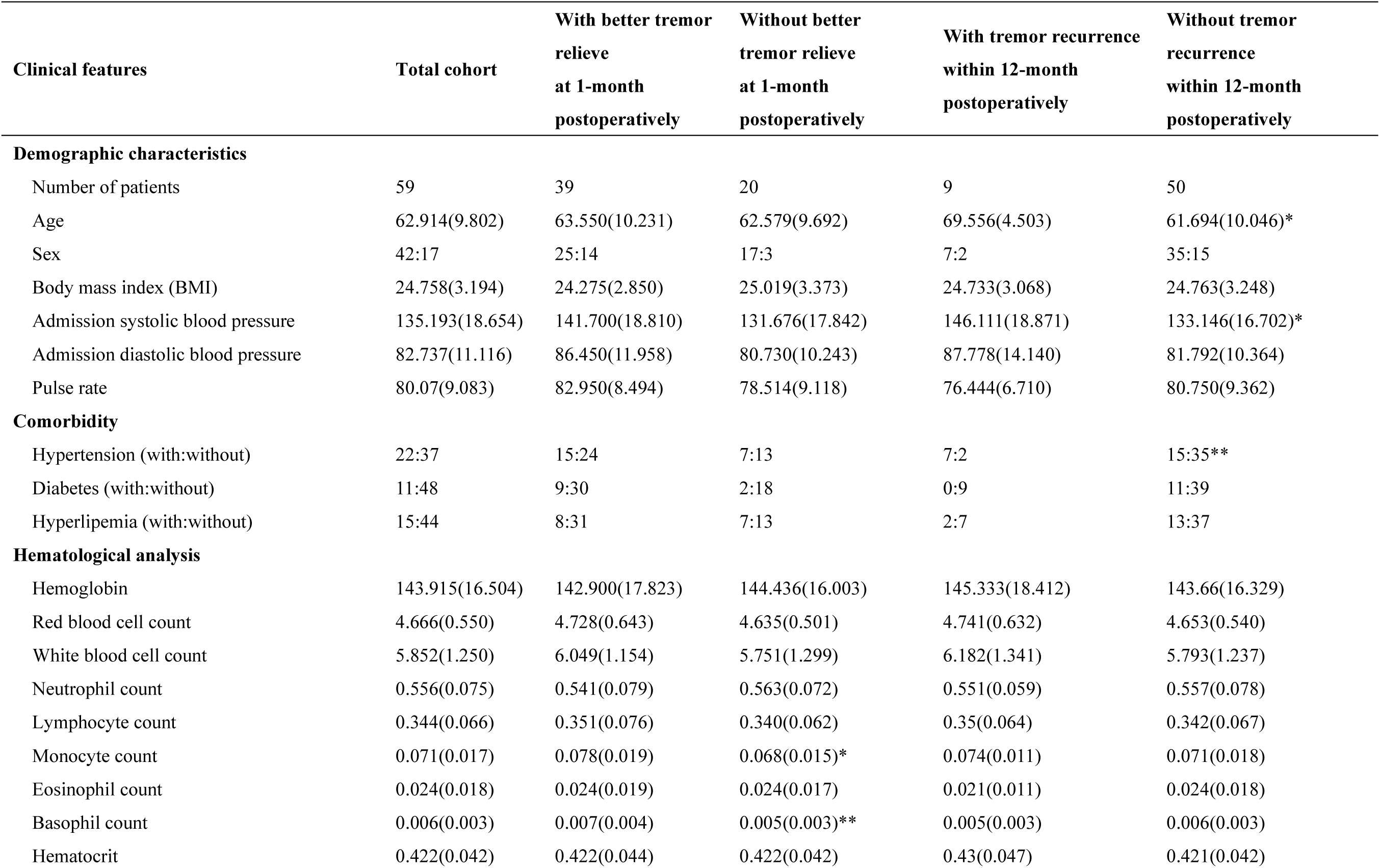
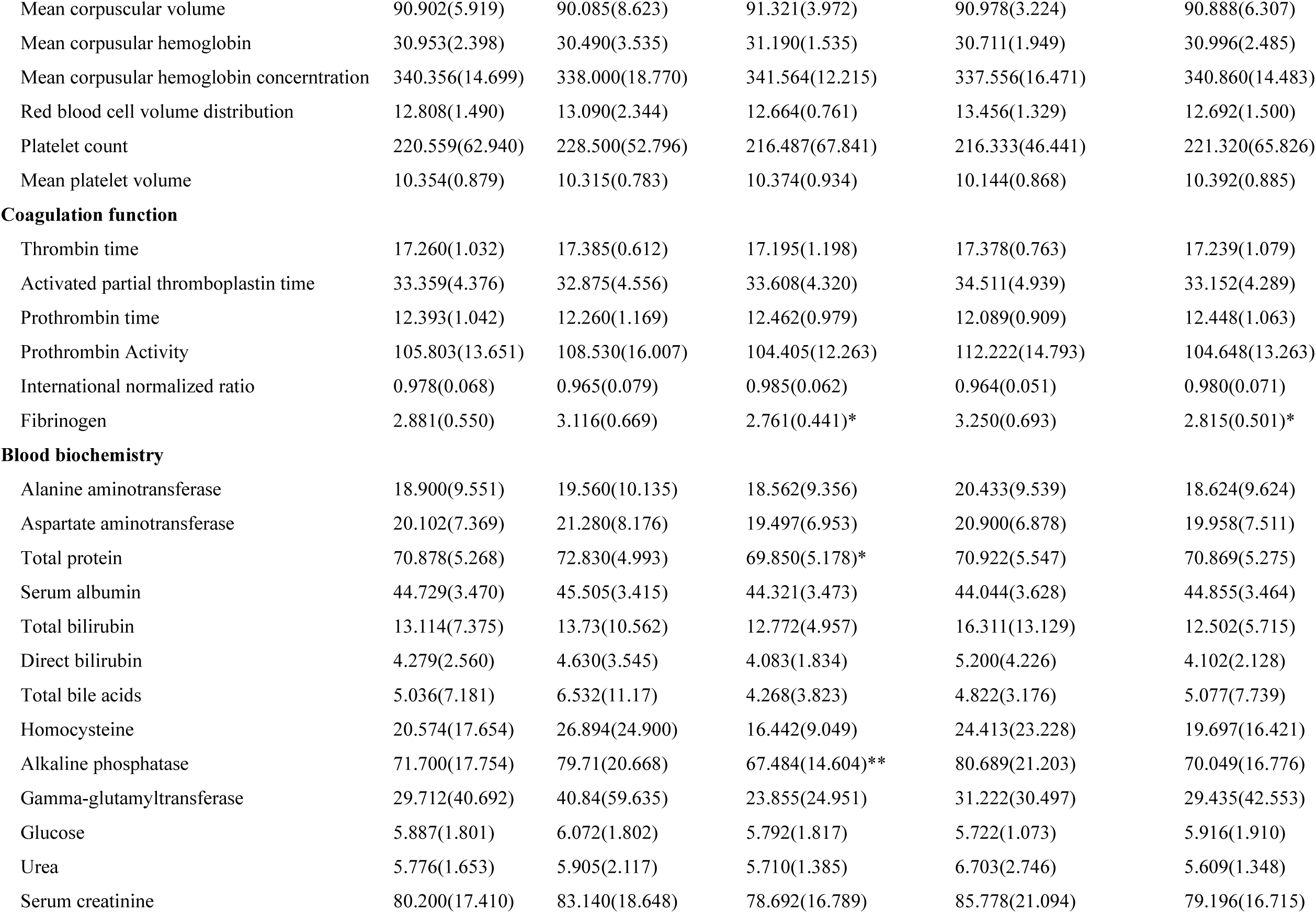
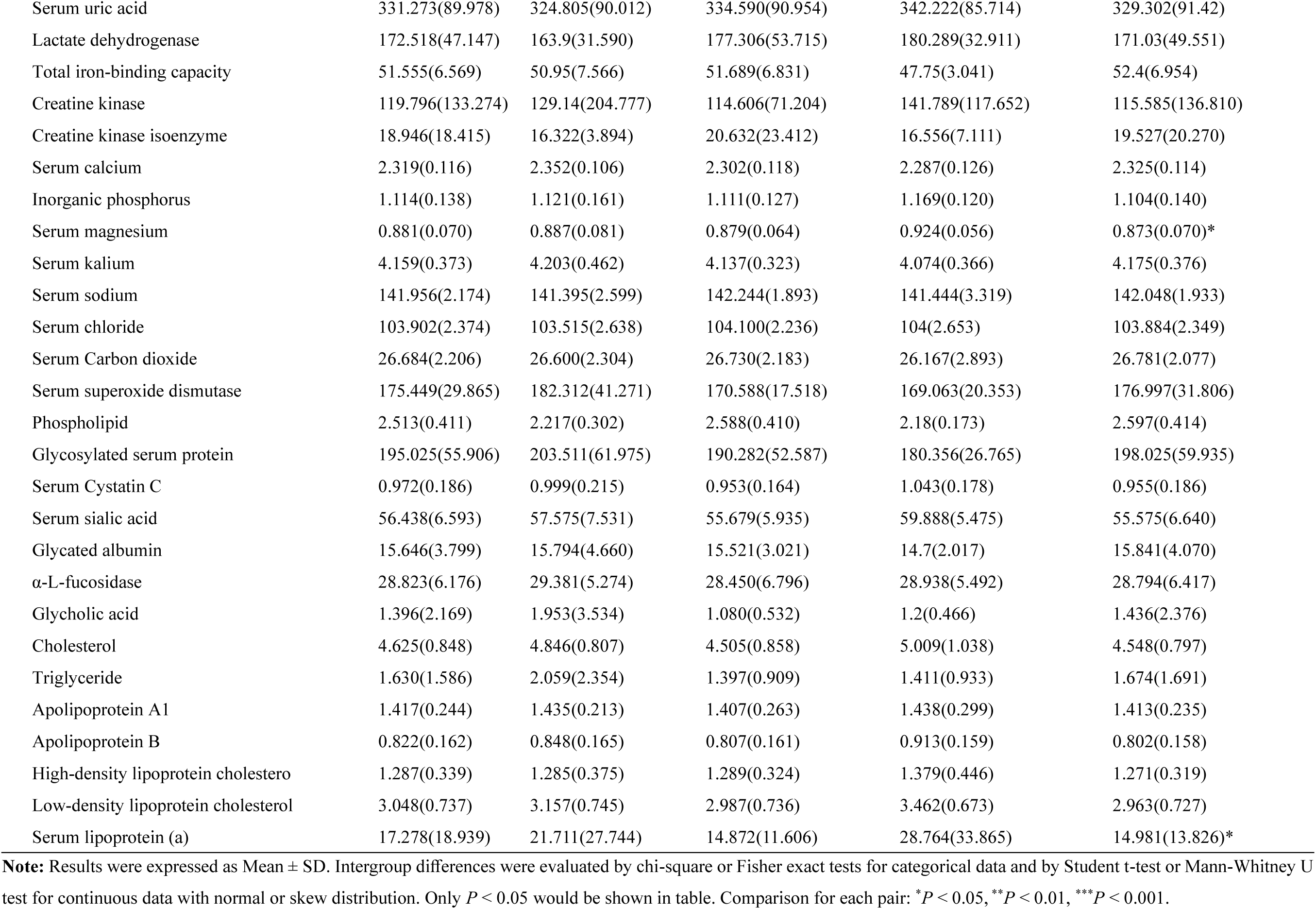
Comparison of demographic and clinical features of ET patients with or without better tremor relieve.

| Clinical features | Total cohort | With better tremor relieve at 1-month postoperatively | Without better tremor relieve at 1-month postoperatively | With tremor recurrence within 12-month postoperatively | Without tremor recurrence within 12-month postoperatively |
| --- | --- | --- | --- | --- | --- |
| **Demographic characteristics** |  |  |  |  |  |
| Number of patients | 59 | 39 | 20 | 9 | 50 |
| Age | 62.914(9.802) | 63.550(10.231) | 62.579(9.692) | 69.556(4.503) | 61.694(10.046)* |
| Sex | 42:17 | 25:14 | 17:3 | 7:2 | 35:15 |
| Body mass index (BMI) | 24.758(3.194) | 24.275(2.850) | 25.019(3.373) | 24.733(3.068) | 24.763(3.248) |
| Admission systolic blood pressure | 135.193(18.654) | 141.700(18.810) | 131.676(17.842) | 146.111(18.871) | 133.146(16.702)* |
| Admission diastolic blood pressure | 82.737(11.116) | 86.450(11.958) | 80.730(10.243) | 87.778(14.140) | 81.792(10.364) |
| Pulse rate | 80.07(9.083) | 82.950(8.494) | 78.514(9.118) | 76.444(6.710) | 80.750(9.362) |
| **Comorbidity** |  |  |  |  |  |
| Hypertension (with:without) | 22:37 | 15:24 | 7:13 | 7:2 | 15:35** |
| Diabetes (with:without) | 11:48 | 9:30 | 2:18 | 0:9 | 11:39 |
| Hyperlipemia (with:without) | 15:44 | 8:31 | 7:13 | 2:7 | 13:37 |
| **Hematological analysis** |  |  |  |  |  |
| Hemoglobin | 143.915(16.504) | 142.900(17.823) | 144.436(16.003) | 145.333(18.412) | 143.66(16.329) |
| Red blood cell count | 4.666(0.550) | 4.728(0.643) | 4.635(0.501) | 4.741(0.632) | 4.653(0.540) |
| White blood cell count | 5.852(1.250) | 6.049(1.154) | 5.751(1.299) | 6.182(1.341) | 5.793(1.237) |
| Neutrophil count | 0.556(0.075) | 0.541(0.079) | 0.563(0.072) | 0.551(0.059) | 0.557(0.078) |
| Lymphocyte count | 0.344(0.066) | 0.351(0.076) | 0.340(0.062) | 0.35(0.064) | 0.342(0.067) |
| Monocyte count | 0.071(0.017) | 0.078(0.019) | 0.068(0.015)* | 0.074(0.011) | 0.071(0.018) |
| Eosinophil count | 0.024(0.018) | 0.024(0.019) | 0.024(0.017) | 0.021(0.011) | 0.024(0.018) |
| Basophil count | 0.006(0.003) | 0.007(0.004) | 0.005(0.003)** | 0.005(0.003) | 0.006(0.003) |
| Hematocrit | 0.422(0.042) | 0.422(0.044) | 0.422(0.042) | 0.43(0.047) | 0.421(0.042) |
| Mean corpuscular volume | 90.902(5.919) | 90.085(8.623) | 91.321(3.972) | 90.978(3.224) | 90.888(6.307) |
| Mean corpuscular hemoglobin | 30.953(2.398) | 30.490(3.535) | 31.190(1.535) | 30.711(1.949) | 30.996(2.485) |
| Mean corpuscular hemoglobin concentration | 340.356(14.699) | 338.000(18.770) | 341.564(12.215) | 337.556(16.471) | 340.860(14.483) |
| Red blood cell volume distribution | 12.808(1.490) | 13.090(2.344) | 12.664(0.761) | 13.456(1.329) | 12.692(1.500) |
| Platelet count | 220.559(62.940) | 228.500(52.796) | 216.487(67.841) | 216.333(46.441) | 221.320(65.826) |
| Mean platelet volume | 10.354(0.879) | 10.315(0.783) | 10.374(0.934) | 10.144(0.868) | 10.392(0.885) |
| **Coagulation function** |  |  |  |  |  |
| Thrombin time | 17.260(1.032) | 17.385(0.612) | 17.195(1.198) | 17.378(0.763) | 17.239(1.079) |
| Activated partial thromboplastin time | 33.359(4.376) | 32.875(4.556) | 33.608(4.320) | 34.511(4.939) | 33.152(4.289) |
| Prothrombin time | 12.393(1.042) | 12.260(1.169) | 12.462(0.979) | 12.089(0.909) | 12.448(1.063) |
| Prothrombin Activity | 105.803(13.651) | 108.530(16.007) | 104.405(12.263) | 112.222(14.793) | 104.648(13.263) |
| International normalized ratio | 0.978(0.068) | 0.965(0.079) | 0.985(0.062) | 0.964(0.051) | 0.980(0.071) |
| Fibrinogen | 2.881(0.550) | 3.116(0.669) | 2.761(0.441)* | 3.250(0.693) | 2.815(0.501)* |
| **Blood biochemistry** |  |  |  |  |  |
| Alanine aminotransferase | 18.900(9.551) | 19.560(10.135) | 18.562(9.356) | 20.433(9.539) | 18.624(9.624) |
| Aspartate aminotransferase | 20.102(7.369) | 21.280(8.176) | 19.497(6.953) | 20.900(6.878) | 19.958(7.511) |
| Total protein | 70.878(5.268) | 72.830(4.993) | 69.850(5.178)* | 70.922(5.547) | 70.869(5.275) |
| Serum albumin | 44.729(3.470) | 45.505(3.415) | 44.321(3.473) | 44.044(3.628) | 44.855(3.464) |
| Total bilirubin | 13.114(7.375) | 13.73(10.562) | 12.772(4.957) | 16.311(13.129) | 12.502(5.715) |
| Direct bilirubin | 4.279(2.560) | 4.630(3.545) | 4.083(1.834) | 5.200(4.226) | 4.102(2.128) |
| Total bile acids | 5.036(7.181) | 6.532(11.17) | 4.268(3.823) | 4.822(3.176) | 5.077(7.739) |
| Homocysteine | 20.574(17.654) | 26.894(24.900) | 16.442(9.049) | 24.413(23.228) | 19.697(16.421) |
| Alkaline phosphatase | 71.700(17.754) | 79.71(20.668) | 67.484(14.604)** | 80.689(21.203) | 70.049(16.776) |
| Gamma-glutamyltransferase | 29.712(40.692) | 40.84(59.635) | 23.855(24.951) | 31.222(30.497) | 29.435(42.553) |
| Glucose | 5.887(1.801) | 6.072(1.802) | 5.792(1.817) | 5.722(1.073) | 5.916(1.910) |
| Urea | 5.776(1.653) | 5.905(2.117) | 5.710(1.385) | 6.703(2.746) | 5.609(1.348) |
| Serum creatinine | 80.200(17.410) | 83.140(18.648) | 78.692(16.789) | 85.778(21.094) | 79.196(16.715) |
| Serum uric acid | 331.273(89.978) | 324.805(90.012) | 334.590(90.954) | 342.222(85.714) | 329.302(91.42) |
| Lactate dehydrogenase | 172.518(47.147) | 163.9(31.590) | 177.306(53.715) | 180.289(32.911) | 171.03(49.551) |
| Total iron-binding capacity | 51.555(6.569) | 50.95(7.566) | 51.689(6.831) | 47.75(3.041) | 52.4(6.954) |
| Creatine kinase | 119.796(133.274) | 129.14(204.777) | 114.606(71.204) | 141.789(117.652) | 115.585(136.810) |
| Creatine kinase isoenzyme | 18.946(18.415) | 16.322(3.894) | 20.632(23.412) | 16.556(7.111) | 19.527(20.270) |
| Serum calcium | 2.319(0.116) | 2.352(0.106) | 2.302(0.118) | 2.287(0.126) | 2.325(0.114) |
| Inorganic phosphorus | 1.114(0.138) | 1.121(0.161) | 1.111(0.127) | 1.169(0.120) | 1.104(0.140) |
| Serum magnesium | 0.881(0.070) | 0.887(0.081) | 0.879(0.064) | 0.924(0.056) | 0.873(0.070)* |
| Serum kalium | 4.159(0.373) | 4.203(0.462) | 4.137(0.323) | 4.074(0.366) | 4.175(0.376) |
| Serum sodium | 141.956(2.174) | 141.395(2.599) | 142.244(1.893) | 141.444(3.319) | 142.048(1.933) |
| Serum chloride | 103.902(2.374) | 103.515(2.638) | 104.100(2.236) | 104(2.653) | 103.884(2.349) |
| Serum Carbon dioxide | 26.684(2.206) | 26.600(2.304) | 26.730(2.183) | 26.167(2.893) | 26.781(2.077) |
| Serum superoxide dismutase | 175.449(29.865) | 182.312(41.271) | 170.588(17.518) | 169.063(20.353) | 176.997(31.806) |
| Phospholipid | 2.513(0.411) | 2.217(0.302) | 2.588(0.410) | 2.18(0.173) | 2.597(0.414) |
| Glycosylated serum protein | 195.025(55.906) | 203.511(61.975) | 190.282(52.587) | 180.356(26.765) | 198.025(59.935) |
| Serum Cystatin C | 0.972(0.186) | 0.999(0.215) | 0.953(0.164) | 1.043(0.178) | 0.955(0.186) |
| Serum sialic acid | 56.438(6.593) | 57.575(7.531) | 55.679(5.935) | 59.888(5.475) | 55.575(6.640) |
| Glycated albumin | 15.646(3.799) | 15.794(4.660) | 15.521(3.021) | 14.7(2.017) | 15.841(4.070) |
| α-L-fucosidase | 28.823(6.176) | 29.381(5.274) | 28.450(6.796) | 28.938(5.492) | 28.794(6.417) |
| Glycholic acid | 1.396(2.169) | 1.953(3.534) | 1.080(0.532) | 1.2(0.466) | 1.436(2.376) |
| Cholesterol | 4.625(0.848) | 4.846(0.807) | 4.505(0.858) | 5.009(1.038) | 4.548(0.797) |
| Triglyceride | 1.630(1.586) | 2.059(2.354) | 1.397(0.909) | 1.411(0.933) | 1.674(1.691) |
| Apolipoprotein A1 | 1.417(0.244) | 1.435(0.213) | 1.407(0.263) | 1.438(0.299) | 1.413(0.235) |
| Apolipoprotein B | 0.822(0.162) | 0.848(0.165) | 0.807(0.161) | 0.913(0.159) | 0.802(0.158) |
| High-density lipoprotein cholestero | 1.287(0.339) | 1.285(0.375) | 1.289(0.324) | 1.379(0.446) | 1.271(0.319) |
| Low-density lipoprotein cholesterol | 3.048(0.737) | 3.157(0.745) | 2.987(0.736) | 3.462(0.673) | 2.963(0.727) |
| Serum lipoprotein (a) | 17.278(18.939) | 21.711(27.744) | 14.872(11.606) | 28.764(33.865) | 14.981(13.826)* |

**Note:** Results were expressed as Mean ± SD. Intergroup differences were evaluated by chi-square or Fisher exact tests for categorical data and by Student t-test or Mann-Whitney U test for continuous data with normal or skew distribution. Only $P < 0.05$ would be shown in table. Comparison for each pair: \* $P < 0.05$ , \*\* $P < 0.01$ , \*\*\* $P < 0.001$ .

### Fibrinogen knock-out and tremor measurement in vivo

Animal care and experimental procedures were in accordance with institutional and NIH protocols and guidelines, and all studies were approved by the Committee on the Ethics of Animal Experiments of Chinese PLA General Hospital. Male C57BL/6 mice aged 6-8 weeks were obtained from the Laboratory Animal Center of Chinese PLA General Hospital. Fibrinogen is composed of pairs of alpha (FGA), beta (FGB) and gamma (FGG) chains, and FGB chain is the rate-limiting factor in the production of the serum fibrinogen hexamer^12^. Male C57BL/6Smoc-Fgb^em1Smoc^ mice aged 6-8 weeks were purchased from Shanghai Model Organisms Center (Cat. NO. NM-KO-200199) as fibrinogen knock-out mice. All mice were maintained under SPF conditions and housed with a 12 h dark/light cycle, humidity of 40-70%, ambient temperature of 20-26 °C and enough supplement of food and water.

Tremor was induced by intraperitoneal injection of harmaline (Sigma-Aldrich, USA). Mice were moved with their home cages to a behavior room and acclimated there for at least one hour before testing. The frequency and intensity of tremor were measured and analyzed using a force-plated accelerometer (Tremor Monitor, San Diego Instruments, USA), which can accurately identify tremor activity from ambulatory/stereotyped movements and global motion activity. Each mouse was placed in the chamber for acclimatization, and then the baseline motion power was recorded for 10 min before administration. Each raw trace of the tremor activity was filtered and converted to power spectra of frequency by a fast Fourier transform on each frame of the experiment using python engine. To quantify the difference in tremor performance of each mouse, the tremor activity ratio was measured as the ratio of the tremor motion power (at tremor frequency bandwidth of 8-15 Hz) over the total motion power (at full motion spectrum of 0-30 Hz) within 10 min duration. The overall tremor activity was expressed by the area under curves (AUCs) derived and was normalized by the averaged baseline. The experiment was performed over five consecutive days.

### Statistical analysis

Most computations were performed in the R engine. Categorical data were presented as number with percentage and continuous data as median with standard error (SD). The linear mixed-effect model (LMM) was used to analyze the trend if necessary. Spearman/Pearson correlation was used to assess the association between clinical variables and tremor symptoms. Intergroup differences were evaluated by chi-square or Fisher exact tests for categorical data and by Student *t*-test or Mann-Whitney U test for continuous data with normal or skew distribution, with False Discovery Rate (FDR) correction if appropriate. Binary logistic regression analyses were performed to identify predictors for therapeutic efficacy or tremor recurrence. Variables with *P* < 0.05 from the univariate regression analysis were included in the subsequent multi-variate analysis and to construct the nomogram. Receiver operating characteristic (ROC) curves were plotted to calculate the AUC, confidence interval by the bootstrap approach to discriminate the performance of the nomogram. In addition, we measured calibration analysis by bootstrap approach and Hosmer-Lemeshow test to assess the agreement of nomogram. A *P* value < 0.05 was used as the criterion for a significant statistical difference.

### Data and code availability

Anonymized data will be made available by request from the corresponding author.

## Results

### Demographic and clinical characteristics

As summarized in Table 1, the mean age of all 59 ET patients was 62.877 ± 9.885 years at the preoperative baseline (Pre-op) and their sex ratios (male:feamle) were 42:17. Among them, average disease duration were 20.741 ± 10.870 years and mean skull density ratio (SDR) were 0.509 ±.113. Of 50 ET patients with family enquire available, 41 of them (82.000%) had a family history of ET. PHQ-9 and MMSE examinations were found in 55 and 37 patients’ records, with scores of 2.655 ± 3.122 and 27.722 ± 1.814 respectively.

There were significant tremor manifestations on the right extremity in 55 ET patients and the left extremity in 4 patients. Their hand tremor scores were 21.810 ± 4.169 and CRST-total scores were 60.517 ± 13.242. We tried to analyze whether the baseline clinical variables were correlated with tremor ratings at baseline, and Spearman correlation analysis identified that multiple measures of coagulation function were correlated with baseline tremor ratings (including International normalized ratio: *P* < 0.05 with hand tremor scores and CRST-A/CRST-B/CRST-total score; fibrinogen: *P* < 0.05 with CRST-A score)(Table 2). However, there was no significant result after FDR verification. Moreover, we also studied the clinical baseline variables based on ET disease duration, and the results showed that patients with disease duration less than 10 years (n = 8) had much higher in homocysteine (*t* = 4.854, *P*_FDR_ = 0.001) and fibrinogen (*t* = 3.357, *P*_FDR_ = 0.049).

**Table 2.** Spearman correlation analysis of baseline clinical variables associated with baseline tremor ratings in ET patients subjected with MRgFUS thalamotomy.

| Baseline tremor ratings/clinical variables | Spearman *r* | Spearman *P* |
| --- | --- | --- |
| **Hand tremor score** |  |  |
| Monocyte count | -0.273 | 0.038 |
| International normalized ratio | 0.297 | 0.023 |
| Glucose | -0.282 | 0.032 |
| Serum superoxide dismutase | -0.390 | 0.013 |
| Phospholipid | -0.525 | 0.045 |
| Serum sialic acid | -0.337 | 0.036 |
| **CRST-A** |  |  |
| Hyperlipemia | -0.358 | 0.008 |
| International normalized ratio | 0.314 | 0.016 |
| Fibrinogen | -0.353 | 0.007 |
| Inorganic phosphorus | 0.261 | 0.050 |
| Monocyte count | 0.414 | 0.001 |
| **CRST-B** |  |  |
| Prothrombin time | 0.361 | 0.005 |
| Prothrombin Activity | -0.275 | 0.036 |
| International normalized ratio | 0.336 | 0.010 |
| **CRST-C** |  |  |
| Age | 0.274 | 0.039 |
| Monocyte count | 0.269 | 0.041 |
| Basophil count | 0.297 | 0.024 |
| Mean corpuscular hemoglobin concentration | -0.272 | 0.039 |
| α-L-fucosidase | -0.357 | 0.026 |
| Glycholic acid | 0.293 | 0.049 |
| **CRST-total** |  |  |
| Hyperlipemia | -0.290 | 0.035 |
| Prothrombin time | 0.368 | 0.004 |
| Prothrombin Activity | -0.262 | 0.047 |
| International normalized ratio | 0.338 | 0.009 |

Note: CRST = Clinical Rating Scale for Tremor. Only the items with $P < 0.05$ in the Spearman correlation analysis were shown.

### Clinical factors for short-term tremor relieve by unilateral MRgFUS thalamotomy

Unilateral MRgFUS thalamotomy resulted in significant tremor relieve (LMM: *P*_for trend_ < 0.001 for hand tremor score and all CRST ratings) (Figure 1A). Their hand tremor scores decreased to 4.368 ± 3.806 points (control ratio = 80.694 ± 15.498%, *P* < 0.001 compared with Pre-op) and CRST-total scores decreased to 20.948 ± 11.657 (control ratio = 66.711 ± 14.639 %, *P* < 0.001 compared with Pre-op) at 1-month postoperatively. All included ET patients presented a good effectiveness to unilateral MRgFUS thalamotomy with the control ratio ranged from 65.196% to 100% at 1-month postoperatively.

**Figure 1.**
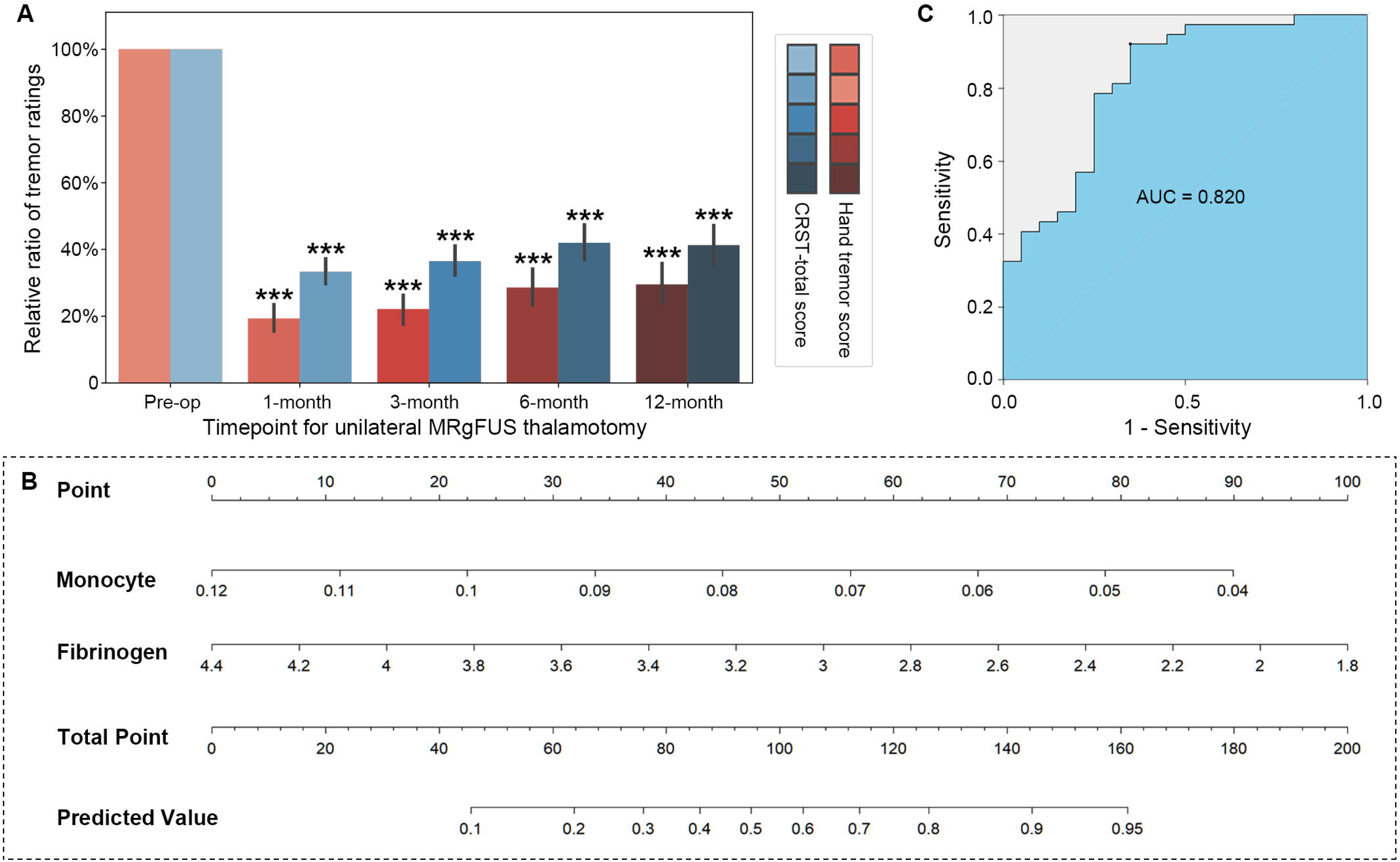
Clinical factors for short-term tremor relieve by unilateral MRgFUS thalamotomy (A) Relative ratio of tremor ratings in ET patients subjected with MRgFUS thalamotomy. (B) Nomogram predicting the better efficiency in tremor relieve (control ratio > 75%) probabilities of ET right after MRgFUS thalamotomy. (C) The ROC curves represent the discrimination ability of the model measured by the AUC is 0.820. (Pre-op = pre-operative; 1-month = 1-month postoperatively; 3-month = 3-month postoperatively; 6-month = 6-month postoperatively; 12-month = 12-month postoperatively. Comparison with Pre-op: \*\*\**P* < 0.001.)

As shown in Table 1, these patients with better short-term tremor relieve (n = 39) had much lower basophil count (*t* = −2.974, *P* = 0.004), lower alkaline phosphatase (*t* = −2.743, *P* = 0.008), lower monocyte count (*t* = −2.312, *P* = 0.025), lower fibrinogen (*t* = −2.309, *P* = 0.025), lower total protein (*t* = −2.275, *P* = 0.027) than those without better tremor relieve (n = 20). All factors with differences were examined using univariate logistic regression model, and it was found that alkaline phosphatase (odds ratio (OR) = 0.956, 95% Confidence Interval (CI) = 0.922-0.992, *P* = 0.016), basophil count (OR = 0, 95% CI = 0-0.001, *P* = 0.026), monocyte count (OR = 0, 95% CI = 0-0.011, *P* = 0.027), fibrinogen (OR = 0.292, 95% CI = 0.096-0.886, *P* = 0.030), total protein (OR = 0.891, 95% CI = 0.794-0.999, *P* = 0.044) were negatively associated short-term tremor relieve efficiency (Table 3). Subsequent multivariate analysis indicated that fibrinogen (OR = 0.182, 95% CI = 0.042-0.796, *P* = 0.024) and monocyte count (OR = 0, 95% CI = 0-0.001, *P* = 0.014) were independently associated with short-term tremor relieve efficiency (Table 3).

**Table 3.**
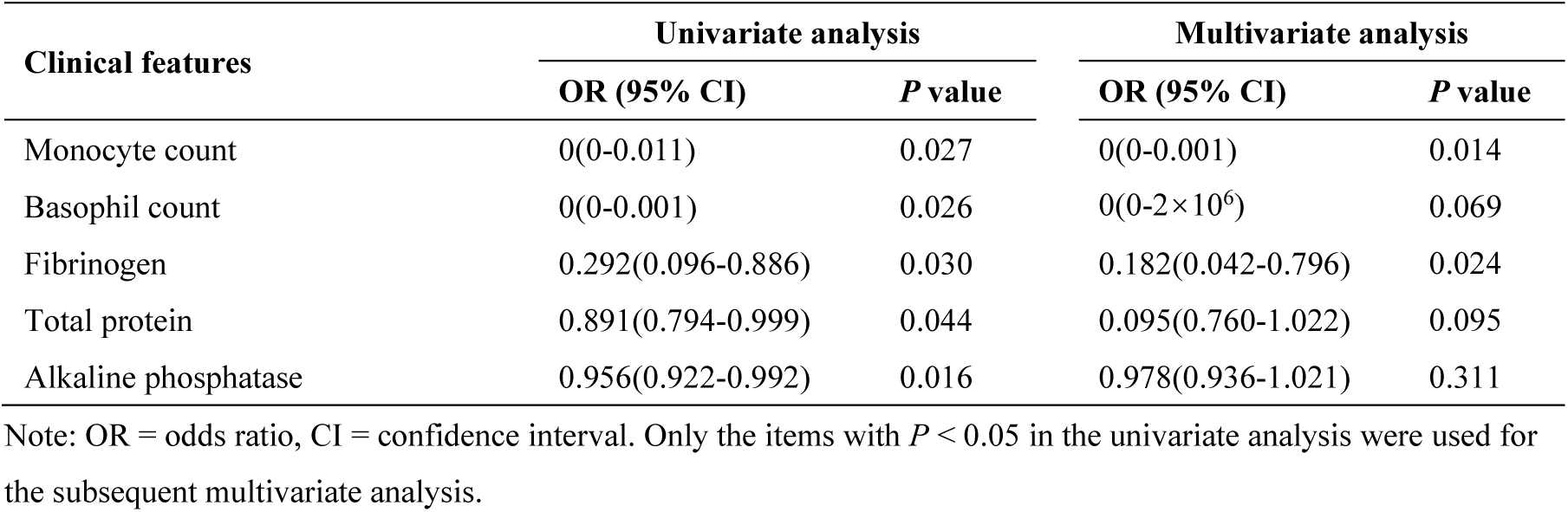
Logistic regression analysis of factors associated with better short-term tremor relieve in ET patients subjected with MRgFUS thalamotomy.

| Clinical features | Univariate analysis |  | Multivariate analysis |  |
| --- | --- | --- | --- | --- |
|  | OR (95% CI) | *P* value | OR (95% CI) | *P* value |
| Monocyte count | 0(0-0.011) | 0.027 | 0(0-0.001) | 0.014 |
| Basophil count | 0(0-0.001) | 0.026 | 0(0-2×10<sup>6</sup>) | 0.069 |
| Fibrinogen | 0.292(0.096-0.886) | 0.030 | 0.182(0.042-0.796) | 0.024 |
| Total protein | 0.891(0.794-0.999) | 0.044 | 0.095(0.760-1.022) | 0.095 |
| Alkaline phosphatase | 0.956(0.922-0.992) | 0.016 | 0.978(0.936-1.021) | 0.311 |

Note: OR = odds ratio, CI = confidence interval. Only the items with *P* < 0.05 in the univariate analysis were used for the subsequent multivariate analysis.

We then used these two factors to establish an individualized prediction nomogram, which can calculate the total point for each ET patient subjected with unilateral MRgFUS thalamotomy and converted it to predicted probabilities of better short-term therapeutic efficiency (Figure 1B). The ability of our nomogram to discriminate the patients with better short-term therapeutic efficiency was excellent as the AUC was 0.820 (95% CI = 0.690-0.919, bootstrap repetition = 1000) (Figure 1C). Besides, the *P*-values of the Hosmer-Lemeshow test is 0.542 and the result of the calibration curve by the bootstrap approach (bootstrap repetition = 1000) indicated our model had a good agreement.

### Clinical factors for long-term tremor recurrence by unilateral MRgFUS thalamotomy

Although it got a good effectiveness to unilateral MRgFUS thalamotomy, a slight decrease in tremor control ratio could be observed at later follow-up of 12 months. Hand tremor scores of ET patients decreased to 6.655 ± 5.842 points (control ratio = 70.484 ± 23.982%, *P* < 0.001 compared with Pre-op) and CRST-total scores decreased to 26.017 ± 17.564 (control ratio = 58.712 ± 23.749%, *P* < 0.001 compared with Pre-op) at 12-month postoperatively. There were three patients experienced significant tremor recurrence at 6-month postoperatively (Figure 2A). A total of nine ET patients (15.254% of 59 patients) finally experienced significant tremor recurrence at 12-month postoperatively, of which one patient almost completely returned to the preoperative state (Figure 2A). We divided these patients into tremor recurrence group.

**Figure 2.**
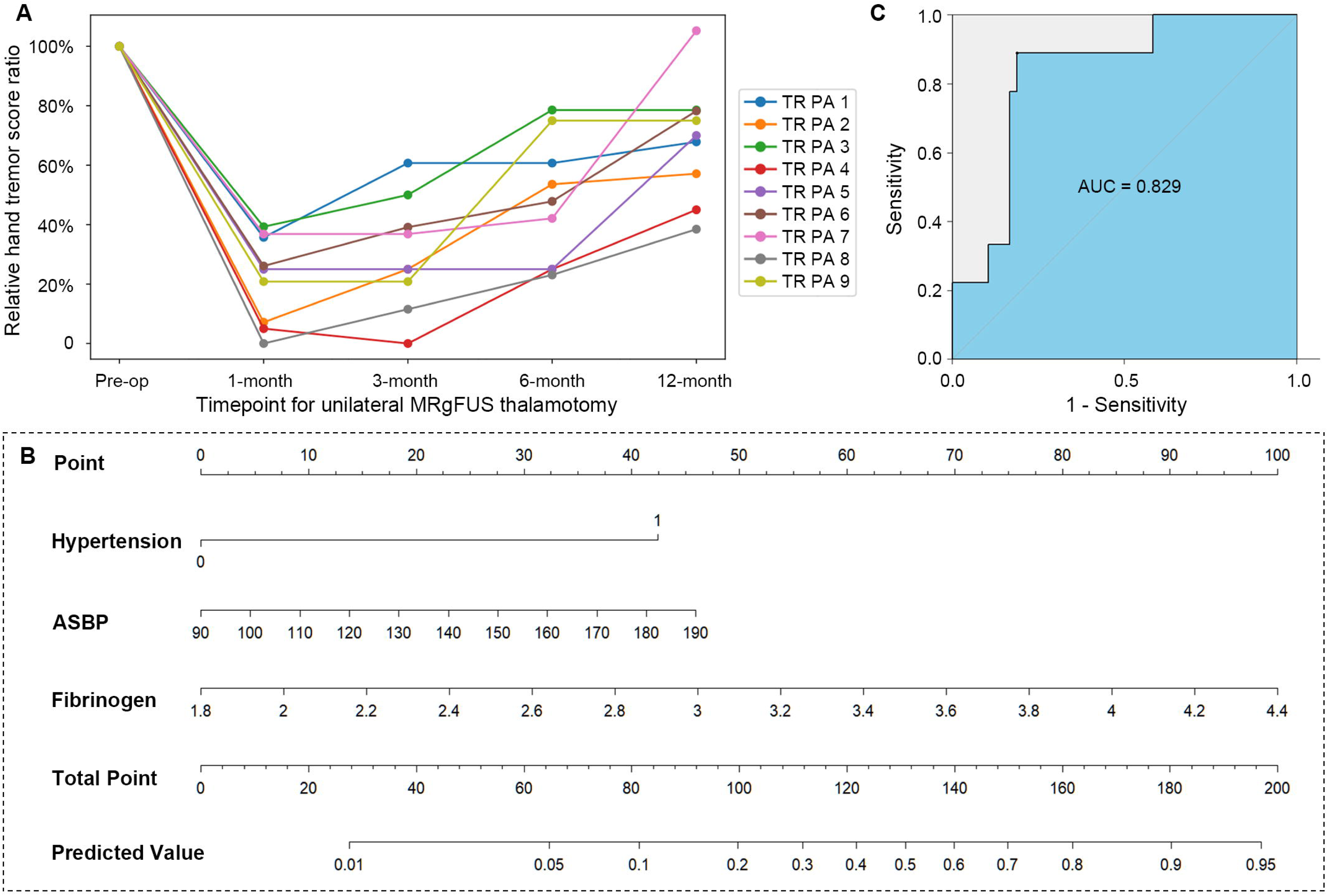
Clinical factors for long-term tremor recurrence by unilateral MRgFUS thalamotomy (A) The temporal change of relative hand tremor control ratio in each ET patient with tremor recurrence within the follow-up of 12 month after MRgFUS thalamotomy. (B) Nomogram predicting the tremor recurrency probabilities of ET patients within the follow-up of 12 month after MRgFUS thalamotomy. (C) The ROC curves represent the discrimination ability of the model measured by the AUC is .829. (Pre-op = pre-operative; 1-month = 1-month postoperatively; 3-month = 3-month postoperatively; 6-month = 6-month postoperatively; 12-month = 12-month postoperatively; ASBP = admission systolic blood pressure.)

It was found that ET patients with tremor recurrence (n = 9) were different from those without tremor recurrence (n = 50) in a younger age (*t* = 2.293, *P* = 0.026), admission systolic blood pressure (*t* = 2.103, *P* = 0.040), a more frequent comorbidity of hypertension (X^2^ = 7.445, *P* = 0.006), higher fibrinogen (*t* = 2.293, *P* = 0.028), higher serum magnesium (*t* = 2.070, *P* = 0.043), higher Lp (a) (*t* = 2.052, *P* = 0.045) than those without tremor recurrence (Table 1). Further univariate analysis identified age (OR = 1.143, 95% CI = 1.012-1.290, *P* = 0.032), admission systolic blood pressure (OR = 1.053, 95% CI = 1.036-1.075, *P* = 0.046), hypertension comorbidity (OR = 7.7, 95% CI = 1.427-41.557, *P* = 0.018), fibrinogen (OR = 4.255, 95% CI = 1.091-16.597, *P* = 0.037), serum magnesium (OR = 6 ×10^5^, 95% CI = 1.076-4×10^11^, *P* = 0.049) were positively associated tremor recurrence (Table 4). Compared to univariate analysis, subsequent multivariate analysis indicated that admission systolic blood pressure (OR = 1.013, 95% CI = 1.010-1.062, *P* = 0.049), hypertension comorbidity (OR = 2.163, 95% CI = 1.412-53.565, *P* = 0.020) and fibrinogen (OR = 1.620, 95% CI = 1.047-24.376, *P* = 0.044) were independent associations with long-term tremor recurrence (Table 4).

**Table 4.** Logistic regression analysis of factors associated with long-term tremor recurrence in ET patients subjected with MRgFUS thalamotomy.

| Clinical features | Univariate analysis |  | Multivariate analysis |  |
| --- | --- | --- | --- | --- |
|  | OR (95% CI) | *P* value | OR (95% CI) | *P* value |
| Age | 1.143(1.012-1.290) | 0.032 | 1.118(0.925-1.351) | 0.250 |
| Admission systolic blood pressure | 1.053(1.036-1.075) | 0.046 | 1.013(1.010-1.062) | 0.049 |
| Hypertension | 7.7(1.427-41.557) | 0.018 | 2.163(1.412-53.565) | 0.020 |
| Fibrinogen | 4.255(1.091-16.597) | 0.037 | 1.620(1.047-24.376) | 0.044 |
| Serum magnesium | $6 \times 10^5$(1.076-$4 \times 10^{11}$) | 0.049 | $6 \times 10^5$(0.016-$2 \times 10^{11}$) | 0.540 |
| Serum lipoprotein (a) | 1.031(0.995-1.067) | 0.097 | - | - |

Note: OR = odds ratio, CI = confidence interval. Only the items with $P < 0.05$ in the univariate analysis were used for the subsequent multivariate analysis.

Therefore, admission systolic blood pressure, hypertension comorbidity and fibrinogen were applied to develop a predictive nomogram for individualized probabilities of long-term tremor recurrence (Figure 2B). It showed that this nomogram could effectively discriminate the ET patients with tremor recurrence with the high AUC of 0.829 (95% CI = 0.965-0.963, bootstrap repetition = 1000) (Figure 2C). Moreover, the *P*-values of the Hosmer-Lemeshow test is .498 and the result of the calibration curve by the bootstrap approach (bootstrap repetition = 1000) indicated this model of long-term tremor recurrence had a good agreement.

### Suppression of fibrinogen knock-out on harmaline-induced tremor in vivo

In above retrospective studies, serum fibrinogen was found not only correlated with tremor symptoms and disease’s duration but also closely associated with tremor relieve and recurrence in ET patients subjected with MRgFUS thalamotomy. We tried to verify the causal role of fibrinogen in the regulation of tremor, a mouse strain of fibrinogen knock-out were introduced in current study.

Both normal mice and fibrinogen knock-out mice were applied for tremor record and measurement by 10 mg/kg harmaline injection. Power spectra indicated harmaline induced a severe action tremor with the peak frequency of 8-15 Hz in normal mice, which was dramatically attenuated in fibrinogen knock-out mice, especially maximum tremor motion power (Figures 3A). The maximum tremor motion power was dramatically attenuated in fibrinogen knock-out mice compared with vehicle-treated mice (*P* < 0.001, Figures 3A). Within 10 min duration, there were significant overall tremor activity in harmaline-treated mice compared with vehicle-treated mice (*P* < 0.001), which was significantly blocked in fibrinogen knock-out mice (*P* < 0.001, Figures 3B).

**Figure 3.**
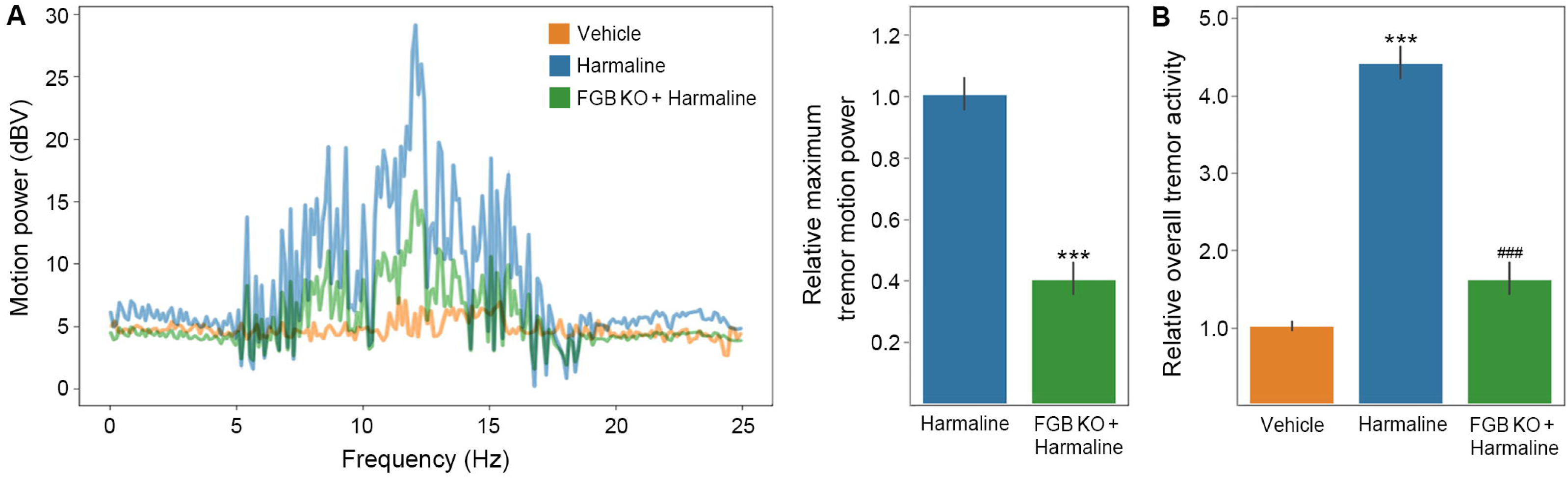
Suppression of fibrinogen knock-out on harmaline-induced tremor in vivo (A) Power spectra for tremor. (FGB KO: in the fibrinogen beta knock-out mice. Comparison with Harmaline: \*\*\**P* < 0.001, n = 12.) (B) Tremor activity. (FGB KO: in the fibrinogen beta knock-out mice. Comparison with Vehicle: \*\*\**P* < 0.001, n = 12. Comparison with Harmaline: ^###^*P* < 0.001, n = 12.)

## Discussion

In current retrospective study, unilateral MRgFUS thalamotomy resulted in significant tremor relieve in ET patients, but significant tremor relapse recurrence has been found in 15.254% patients within the follow-up of 12 months. We conducted a thorough profiling of their baseline clinical features for risk factors, and the evidence regarding fibrinogen-associated differences in the ET disease duration, short-term therapeutic efficiency and long-term tremor relapse of unilateral MRgFUS thalamotomy were provided. Nomogram models of MRgFUS thalamotomy prognosis based on baseline clinical factors was constructed, and they have good predictive power and ability for tremor outcomes. Additional *in vivo* study identified fibrinogen knock-out promoted a great suppression on ET mouse model induced by harmaline.

As an “incisionless” treatment, the application of MRgFUS thalamotomy has experienced quick growth in recent years for medically refractory ET and tremor-dominant PD. A growing accompanying criticism is about its variety of different efficacies (tremor improve rate ranging from 43% to 83%), with loss in efficacy and of tremor recurrence in long-term prognosis^2–4^. However, there still lack a wide accepted criteria for the effectiveness and tremor recurrence for MRgFUS thalamotomy. Some studies use a 5-point increase in Fahn-Tolosa-Marin rating as a measure of tremor recurrence^13^, while others defined as loss of >33% of the efficacy at 3 months relative to a short time after the treatment^5^. Establishment of the consensus as soon as possible is of great significance to further optimize its application.

The key inspiration from this study was a vital role of fibrinogen in the occurrence and development of tremor diseases. Although there are no reports related to fibrinogen in the literature search of clinical studies on ET until now, multiple studies have found that fibrinogen is closely involved in the pathophysiological mechanism of PD: (1) A longitudinal prospective study of 8,006 Japanese-American men has identified 61 prevalent cases and 61 incident cases of PD during the follow-up with high fibrinogen level after adjusting^14^. (2) Cross-sectional study in 40 PD patients has suggested that an abnormal aggregation of fibrinogen and the formation of fibrin amyloid in CSF, suggesting unusual coagulation phenomenon as well as systemic inflammation^15^. (3) Autopsies of 12 PD patients have also revealed significant perivascular deposition of fibrinogen in the striatum^16^, as well as elevated levels of palmitoylated fibrinogen^17^. (4) Additionally, a controlled clinical study investigating 71 patients with carpal tunnel syndrome observed co-deposition and interaction between fibrinogen and α-synuclein^18^. As so far, various studies strongly suggest the close relationship between ET and PD in the clinical manifestations (18.8% of ET patients also sustained rest tremor and 24.2% of them had Lewy bodies in the brainstem^19,20^) and in the high risk of disease course (up to 20% ET patients will go on to develop PD^21^). The data of nine PD patients as previously studied has been introduced for cross-prediction test^10^. The result suggested a generalization of the above nomogram models in PD patients to some extent (Supplementary result 1). However, these cross-prediction results can only be used as a limited and preliminary reference, and more reliable results require more studies in ET and PD patients to better figure out the potential role of fibrinogen in their pathogenesis and clinical features.

Furthermore, recent researchers are beginning to emphasize unique structure of fibrinogen, which contains multiple binding sites for receptors and proteins, resulting in a pluralistic signaling network and pleiotropic functions in neurodegeneration^22^. For example, fibrinogen induces initial neuroinflammatory via CD11b/CD18 on microglia^23^, hinder neurite outgrowth by acting as a ligand for the β3 integrin receptor on neurons^24^, and stimulates the generation of reactive oxygen species and mitochondrial superoxide^25^. Phalguni et al. found intraperitoneal injecting fibrinogen into mice for 48 hours would induce PD-like behaviors such as slow movement and memory deficit, and their further pathological studies found that mice showed loss of dopaminergic neurons in the substantia nigra, compensatory hypertrophy of neurons, and decreased expression of tyrosine hydroxylase in the striatum^26^. Moreover, recent work of Gavin et al. identified the transcript expression of FGA, FGB and FGG in neurons and astrocytes, suggesting the extensive involvement of fibrinogen in various pathophysiological processes of the central nervous system^27^. In our analysis of Allen atlas, we found that intrinsic expression of fibrinogen beta chain was highly concentrated in the cerebellum of human and mouse, which is well recognized as the major neurodegenerative site in the pathogenesis of ET. Therefore, how fibrinogen is involved in the regulation of ET tremor still needs further in-depth study to confirm.

### Limitations

Some limitations of our study should be mentioned. First, the single-center retrospective design and the small sample size are major limitations of the study. The retrospective nature of our present study just only prompted an association not a causal link. The small sample size also has introduced the risk of bias to affect the results of multivariate regression analysis and limit the power to detect associations. Second, although use of multivariate analysis might minimize confounding bias, potential confounders may still exist due to unidentified variables due to the lack of a control group (such as drug treatment). Third, lack of clinical examination at the follow-up timepoints and not considering drug use may lead to extra biased interpretation for current results. Last, the follow-up period of 12 months may not be long enough to capture the long-term durability of the identified predictors. Therefore, the current study should be taken as preliminary evidence that needs to be further validated in larger prospective and multicenter cohort studies with sufficient clinical records.

## Conclusions

Our study demonstrated that clinical baseline variables (especially fibrinogen) are associated with prognostic differences (short-term tremor relieve and long-term tremor recurrence) in patients treated with MRgFUS thalamotomy. Preoperative evaluation of clinical factors has important predictive significance for short - and long-term postoperative outcomes for patients treated with MRgFUS thalamotomy.

## Data Availability

Anonymized data will be made available by request from the corresponding author.

## Acknowledgements

None

