## Supplementary Material for "Fibrinogen is the key factor associated with tremor relieve and recurrence in the ET patients subjected with unilateral MRgFUS thalamotomy"

**Supplementary result 1**

### 1. Supplementary Methods

#### *1.1 Study population of unilateral MRgFUS thalamotomy*

As unilateral MRgFUS thalamotomy is also widely used for tremor control in patients with tremor-dominant PD, we tried to test the generalizability of the ET-based nomogram models to PD patients subjected with unilateral MRgFUS thalamotomy.

We developed and validated a predictive model in a single-center cohort study (NCT04570046 registered at clinicaltrials.gov) of patients with unilateral MRgFUS thalamotomy between January 2018 to September 2020. We developed and validated a predictive model in a single-center cohort study (NCT04570046 registered at clinicaltrials.gov) of patients with unilateral MRgFUS thalamotomy between January 2018 to September 2020. The observational data of all consecutive patients were retrospectively collected in Chinese PLA General Hospital**.** It was approved by the institutional review board and the local scientific advisory committee. The data were anonymous, and the requirement for informed consent was therefore waived. The work has been reported in line with the STROCSS criteria[1].

Participants who fulfilled the following criteria were included: 1) admitted fulfilling diagnostic criteria for tremor-dominant PD; 2) unilateral tremor manifestations on one extremity; 3) received unilateral MRgFUS thalamotomy. Unilateral MRgFUS thalamotomy in contralateral Vim was performed in a 3T MRI suite (Discovery 750, GE Healthcare, USA) using the ExAblate Neuro focused ultrasound system (InSightec, Israel) as previous reports [10]. Participants with one or more of the following conditions were excluded: 1) missing tremor assessment data at 1 or less than 3 tremor assessments made within 12 months of follow-up; 2) missing values in clinical baseline variables at hospitalization; 3) coexistence of other major diseases; and 4) undergone other brain surgeries. Finally, of the final eligible patients, 9 were tremor-dominant PD (Dataset 2).

### 2. Results

#### *2.1 Cross prediction of treatment outcome in eligible PD patients subjected with unilateral MRgFUS thalamotomy*

The hand tremor data and demographic characteristics of these 9 PD patients are consistent with that we've been reporting[2]. Tremor performance of these PD patients was evaluated pre-operatively and 1/3/12-month postoperatively.

As for short-term tremor relieve, there was one PD patient had a bad therapeutic effectiveness to MRgFUS thalamotomy with control ratio of hand tremor score < 50% (16.7%) at 1-month postoperatively. Including this one, a total of two PD patients did not have better therapeutic efficiencies after unilateral MRgFUS thalamotomy (no higher than 75%: 16.7% and 62.5% respectively), and they also got the lowest two predicted values based on our individualized prediction nomogram of better short-term tremor relieve (77% and 81% *vs.* 82.5%, 83%, 86%, 91.5%, 92%, 100%, 100% for probabilities).

Moreover, with the exception of the patient with bad effectiveness, the remaining eight PD patients did not have significant tremor recurrence within 12-month. They all scored very low predicted values at the nomogram model of tremor recurrence (2%, 3%, 4%, 4%, 8%, 19%, 25%, 27% for probabilities).

### Supplementary result 2

### 1. Supplementary Methods

#### *1.1 Allen brain atlas for transcript distributions*

Neuroanatomically precise, genome-wide maps of transcript distributions are critical resources to study genomic genetic architecture. Allen Institute for Brain Science offer the first anatomically and genomically comprehensive three-dimensional human brain map constructed by the brains of six human donors with no history of psychiatric or neuropathological disorders (referred as Allen Human Brain Atlas (AHBA)) [3], as well as mice brain map (referred as Allen Mouse Brain Atlas (AMBA))[4]. We extracted human brain gene transcriptome data were from the AHBA, which contains a database of 20,737 gene expression levels represented by 58,692 probes. All these gene expressions were matched for 274 ROIs of the Brainnetome atlas using the toolbox Abagen[5]. The microarray expression data were combined across donors, and the representative expressions were calculated for each brain regions, including cerebral cortex (n = 218), basal ganglia (n = 12), thalamus (n = 16), cerebellum (n = 27).

We also extracted in-situ hybridization (ISH) data for fibrinogen-related transcripts, including fibrinogen alpha (FGA) (https://mouse.brain-map.org/experiment/show/68442907), fibrinogen beta (FGB) (https://mouse.brain-map.org/experiment/show/68638095), and fibrinogen gamma (FGG) (https://mouse.brain-map.org/experiment/show/576438). The detail experiment procedure and antisense probe information could be found in above sites.

### 2. Results

#### *2.1 Fibrinogen beta is highly expressed in cerebellum*

We matched the three major gene expressions of fibrinogen (FGA, FGB and FGG) from the AHBA microarray for the Brainnetome atlas. It showed that, in the human brain, the expressions of FGA and FGG were far from intensity-based filtering of 0.3, while FGB obtained a high expression, especially in the cerebellum (cerebellum *vs.* cerebral cortex: $P < 0.001$; cerebellum *vs*. basal ganglia: $P < 0.001$; cerebellum *vs*. thalamus: $P < 0.05$) (Figure S1A). To further verify this result, we collected ISH data from AMBA mice brain. The staining also showed that only FGB was highly expressed in the mice brain, also especially in the cerebellum (Figure S1B).

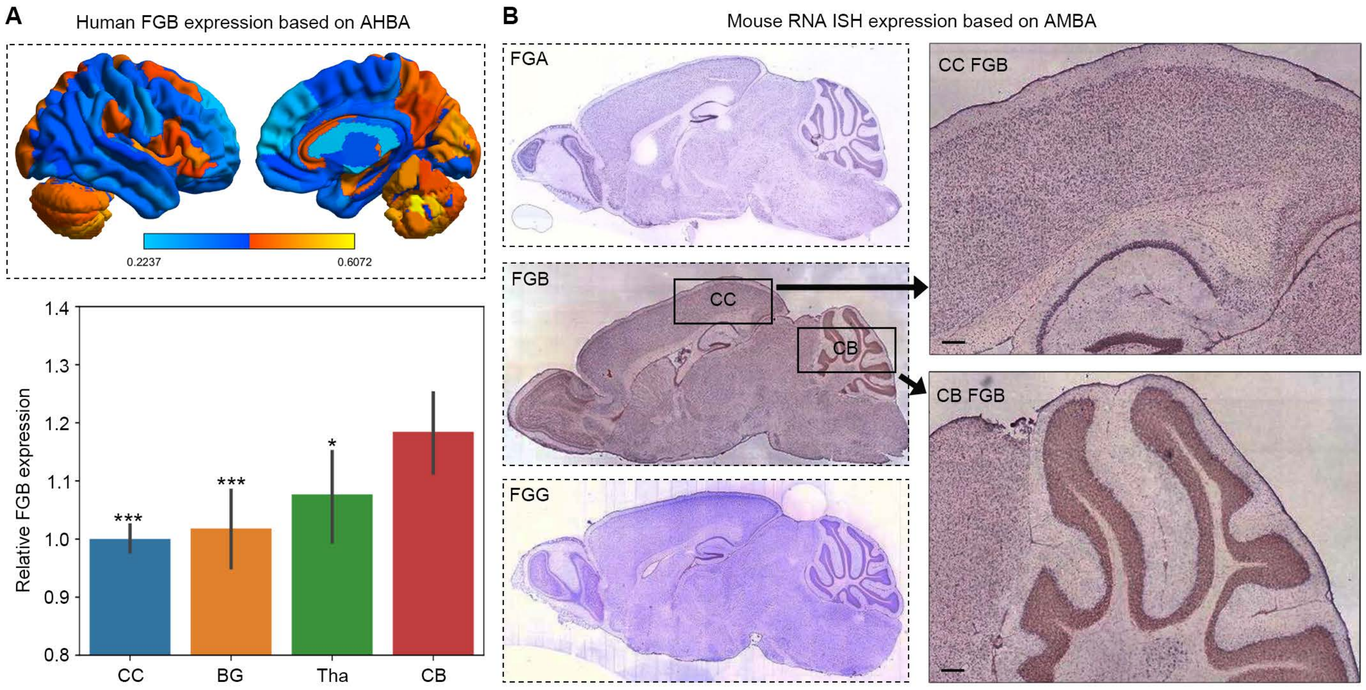

A
Human FGB expression based on AHBA
0.2237
0.6072
1.4
1.3
1.2
1.1
1.0
0.9
0.8
Relative FGB expression
***
***
*
CC
BG
Tha
CB
B
Mouse RNA ISH expression based on AMBA
FGA
FGB
CC
CB
FGG
CC FGB
CB FGB

**Figure S1 Suppression of Fibrinogen beta knock-out on harmaline-induced tremor *in vivo***

**(A)** Human FGB expression based on AHBA. (CC: cerebral cortex, BG: basal ganglia, Tha: thalamus, CB: cerebellum. Comparison with CB: $^{*}P < 0.05$, $^{***}P < 0.001$) **(B)** Mouse RNA ISH expression based on AMBA.
